# Comprehensive Pathogen Profiling in Adult Patients with Severe Acute Respiratory Infections Using Metagenomic Next-Generation Sequencing of Sputum Samples

**DOI:** 10.64898/2025.12.12.25342137

**Authors:** Nang Kham-Kjing, Rathakarn Kawila, Patcharaporn Tariyo, Kittiyaporn Puapun, Nicole Ngo-Giang-Huong, Sayamon Hongjaisee, Woottichai Khamduang

**Affiliations:** Department of Medical Technology, Faculty of Associated Medical Sciences, Chiang Mai University, Chiang Mai, Thailand; LUCENT International Collaboration, Faculty of Associated Medical Sciences, Chiang Mai University, Chiang Mai, Thailand; Nakornping Hospital, Chiang Mai, Thailand; Maladies Infectieuses et Vecteurs: Écologie, Génétique, Évolution et Contrôle (MIVEGEC), Agropolis University Montpellier, Centre National de la Recherche Scientifique (CNRS), Institut de Recherche Pour le Développement (IRD), Montpellier, France; LMI PRESTO, Faculty of Associated Medical Sciences, Chiang Mai University, Chiang Mai, Thailand; Research Institute for Health Sciences, Chiang Mai University, Chiang Mai, Thailand

**Keywords:** SARI, mNGS, long-read sequencing, metagenomic sequencing, ONT, nanopore sequencing

## Abstract

Severe Acute Respiratory Infection (SARI) remains a major global health burden, yet conventional diagnostics frequently fail to identify the causative pathogens. This study aimed to comprehensively characterize the respiratory microbial landscape in adult SARI and to evaluate the clinical utility of an optimized long-read metagenomic next-generation sequencing (mNGS) workflow. A total of 101 respiratory specimens from hospitalized adults in northern Thailand (November 2023 to April 2024) were analyzed using SMART-9N–based cDNA synthesis, Oxford Nanopore sequencing, and a streamlined bioinformatics pipeline. Integrated clinical and laboratory data were used to assess associations with disease severity. Long-read mNGS demonstrated superior diagnostic yield compared to multiplex PCR and culture, detecting pathogens in 78% of cases and uniquely identifying etiologic agents in 18% of specimens negative by routine diagnostics, including *Klebsiella pneumoniae*, *Haemophilus parainfluenzae*, and SARS-CoV-2. High concordance was observed for respiratory viruses, with genome-wide coverage supporting accurate detection. Phylogenetic analysis revealed the co-circulation of HRV-A, HRV-B, and HRV-C, with HRV-C predominating. Multivariable analysis identified male sex and reduced oxygen saturation as independent predictors of severe disease. These findings underscore the diagnostic power of long-read mNGS for uncovering atypical, unculturable, and polymicrobial infections in adult SARI and support its integration into respiratory pathogen surveillance.

## Introduction

Acute respiratory infections (ARIs) remain a leading cause of hospitalization and mortality globally, with pneumonia accounting for a substantial proportion of respiratory-related deaths (1). Severe Acute Respiratory Infection (SARI), as defined by the World Health Organization (WHO), is characterized by an acute respiratory illness with a history of fever or a measured temperature ≥38°C, accompanied by cough within 10 days of symptom onset, and necessitating hospitalization (2). Representing the severe spectrum of respiratory infections, SARI often imposes a considerable burden on healthcare systems (3). Despite its clinical significance, the causative pathogen remains unidentified in a substantial proportion of SARI cases when relying on conventional diagnostic methods (4).

Although most SARI cases are attributed to respiratory viruses, bacterial and fungal pathogens can also play etiological roles, often as part of polymicrobial infections. While multiplex PCR assays have enhanced sensitivity for selected respiratory pathogens and allow simultaneous detection of multiple agents, they are inherently limited by their targeted nature, failing to identify unexpected, atypical, or emerging organisms. In contrast, metagenomic next-generation sequencing (mNGS) provides a comprehensive and unbiased approach for pathogen detection, capable of identifying both known and novel organisms without prior assumptions (5, 6). A recent study has demonstrated that mNGS can detect unculturable or unanticipated organisms frequently missed by conventional diagnostics (7), offering new insights into pathogen ecology in SARI.

Recent findings suggest that SARI patients often harbor multiple pathogens concurrently (8), with some organisms potentially exacerbating disease severity. However, the relationship between microbial diversity and clinical outcomes remains unclear. One hypothesis posits that higher microbial diversity may reflect synergistic pathogenicity contributing to severe disease (9, 10), whereas another suggests it may indicate the absence of a dominant pathogen, correlating with milder illness (11).

Beyond microbial factors, host characteristics such as age, comorbidities, and immune status are also critical determinants of disease severity and pathogen distribution. For example, infants and young children are frequently affected by viruses like respiratory syncytial virus (RSV) or influenza. (12), whereas older adults tend to have distinct pathogen profiles and face greater risks due to underlying conditions (13, 14). Understanding how these host factors influence pathogen distribution may elucidate the heterogeneity in clinical outcomes among SARI patients. Although multiple studies have explored the etiology of SARI, the majority have relied on targeted PCR-based diagnostics (12, 15, 16), which may overlook clinically relevant pathogens. There is limited research employing metagenomic sequencing to comprehensively profile both common and uncommon respiratory pathogens in SARI. Furthermore, the interaction between pathogen diversity and clinical severity remains poorly characterized. Addressing these knowledge gaps is essential for refining diagnostic approaches and improving our understanding of pathogen-associated outcomes.

In this retrospective study, we integrate mNGS data with clinical and outcome variables to provide a high-resolution characterization of the SARI etiological landscape. We aim to evaluate the diagnostic utility of mNGS for detecting pathogens missed by routine diagnostics and to determine whether specific microbial profiles or host factors correlate with disease severity. Our findings may inform clinical decision-making, guide antimicrobial stewardship, and enhance pathogen surveillance strategies for adult SARI populations.

## Materia and methods

### Study design and patient enrollment

This retrospective study analyzed adult patients hospitalized with Severe Acute Respiratory Infection (SARI) at a regional hospital in Chiang Mai, northern Thailand, between November 2023 and April 2024. Inclusion criteria were based on the World Health Organization (WHO) case definition for SARI, which includes acute respiratory illness with onset of fever (≥38°C) and cough within 10 days, requiring hospital admission. A total of 101 adult patients met these criteria and were included in the analysis. Clinical and laboratory data were retrieved from hospital records.

### Sample collection and nucleic acid extraction

Remnant respiratory specimens collected during routine diagnostic care were used for analysis. Sputum samples were treated with an equal volume of 0.1% dithiothreitol, DTT, (Thermo Fisher Scientific, MA, USA) and incubated at room temperature until complete liquefaction to reduce viscosity and homogenize the specimen. The liquefied samples were centrifuged at 10,000×g for 3 minutes to pellet debris, and the supernatant was collected for nucleic acid extraction. For the nasopharyngeal swab sample, no pretreatment step was applied. Total nucleic acids were extracted from 200 µL of each liquefied sputum or nasopharyngeal sample using the QIAamp® Viral RNA Mini Kit (Qiagen GmbH, Hilden, Germany) following the manufacturer’s protocol.

### Host DNA depletion

An optimized metagenomic next-generation sequencing (mNGS) workflow developed in our previous study was employed (17). To deplete host DNA, extracted nucleic acids were treated with RNase-Free DNase Set (Qiagen GmbH, Hilden, Germany) at room temperature, following the manufacturer’s instructions. The treated nucleic acids were then purified using the RNA Clean & Concentrator™-25 Kit (Zymo Research, CA, USA).

### mNGS library preparation and Oxford Nanopore sequencing

Complementary DNA (cDNA) was synthesized using SMART-9N primers (**S**witching **M**echanism **a**t the 5′ end of **R**NA **T**emplate with random **9-n**ucleotide primers), following our previously described protocol (17). Briefly, a 12 µL reaction contained 0.16 µM 9N primer, 0.83 mM dNTPs, and purified nucleic acids, incubated at 65°C for 5 minutes. First-strand synthesis was performed by combining the annealed RNA with 8 µL of a mixture containing 2.5× SuperScript™ IV buffer, 12.5 mM DTT, 5 U RNase inhibitor, 25 U SuperScript™ IV reverse transcriptase (Thermo Fisher Scientific, MA, USA), and 0.25 µM strand-switching primer. The reaction was then incubated at 42°C for 90 minutes, followed by 70°C for 10 minutes. Amplification of cDNA was carried out in 25 µL of reaction containing 1× Q5® reaction buffer, 0.2 µM dNTPs, 0.8 µM PCR primer (AAGCAGTGGTATCAACGCAGAGT), 0.02 U of Q5® High-Fidelity DNA Polymerase (New England Biolabs, MA, USA), and 2.5 µL of cDNA. PCR cycling included an initial denaturation at 98°C for 45 seconds; 30 cycles of 98°C for 15 seconds, 62°C for 15 seconds, and 65°C for 5 minutes; with a final extension at 65°C for 10 minutes. The amplified products were purified using magnetic bead cleanup, VAHTS DNA Clean Beads (Vazyme, Nanjing, China) in equal ratio, and quantified using the DeNovix dsDNA Broad Range Assay Kit (DeNovix Inc., DE, USA) to ensure sufficient yield for sequencing. Sequencing libraries were prepared with the Oxford Nanopore Native Barcoding Kit (ONT-SQK-NBD114.24, Oxford Nanopore Technologies, Oxford, UK) following the manufacturer’s protocol. Unique barcodes were ligated to each sample to allow multiplexing up to 12 samples per flow cell. Barcoded libraries from multiple samples were pooled in equimolar amounts and subjected to adapter ligation. The prepared library was quantified and loaded onto a MinION flow cell (R10.4.1, FLO-MIN114, Oxford Nanopore Technologies, Oxford, UK) for sequencing using the ONT MinION Mk1C platform (Oxford Nanopore Technologies, Oxford, UK). Sequencing was conducted for up to 72 hours per flow cell.

### Bioinformatics analysis

Bioinformatic analysis followed protocols established in our prior study (17). Briefly, raw sequencing data were basecalled using the Dorado basecaller with a high-accuracy model via the MinKNOW software (v24.11.10). Reads that passed the quality filter (minimum average Q-score of 9) were retained and demultiplexed by barcode; simultaneous adapter trimming was performed during this step. Summary sequencing metrics were generated and inspected using NanoComp v1.23.1, and read length and quality distributions were visualized with NanoPlot v1.42.0 for quality control (18). To remove any residual primer or adapter sequences introduced by the SMART-9N protocol, we trimmed reads for the known adapter/primer overhangs using Cutadapt v5.0 (19). We further filtered the reads with fastp v0.24 to discard reads shorter than 200 base pairs (bp), ensuring that only high-quality, longer reads were retained for downstream analysis (20). Host-derived sequences were then computationally subtracted by mapping the reads to the human reference genome (GRCh38) using Minimap2 v2.28-r1209 with parameters optimized for nanopore reads (-x map-ont) (21). Reads aligning to the human genome were identified and removed, and only the unmapped (putatively non-human) reads were kept for downstream microbial analysis. The host-depleted reads were subjected to de novo assembly to reconstruct genomes of detected organisms. Assembly was performed using MetaFlye v2.9.5-b1801 with the “--nano-hq” parameter to leverage the high-quality long reads (22). The assembly outputs were then polished to improve base-level accuracy. We used the Medaka tool v2.0.1 to perform consensus polishing on the assembled contigs, which corrects sequencing errors by realigning the raw reads to the contigs and refining the consensus (23). For taxonomic identification, the assembled contigs were analyzed using Kraken2 v2.1.3 with the Kraken “PlusPF” database, which includes the full RefSeq bacterial, viral, archaeal, protozoal, and fungal sequences (24). We visualized the composition of each sample’s metagenome using KronaTools v2.8.1 to create interactive hierarchical pie charts of taxonomic relative abundance (25). To access the genome coverage of the potential pathogens, we mapped the host-filtered reads to the respective reference genomes of key pathogens using Minimap2 and calculated per-base coverage depth using SAMtools v1.21 (26).

### Phylogenetic analysis

For phylogenetic tree reconstruction, HRV-positive samples from this study were compared with reference prototypes downloaded from the GenBank database. All consensus sequences were combined and aligned using MAFFT (v7.526) with the --auto strategy (27). Poorly aligned positions and high-gap columns were removed using trimAl (v1.5.rev0) with the automated1 algorithm to retain reliably aligned regions (28). Maximum-likelihood phylogenetic trees were reconstructed using IQ-TREE2 (v2.4.0), applying the ModelFinder Plus (MFP) algorithm for best-fit substitution model selection, and branch support was assessed using 1,000 ultrafast bootstrap replicates (UFBoot2) together with 1,000 SH-aLRT tests (29–31). Enterovirus D68 (EV-D68) reference sequences were included as an outgroup to root the HRV phylogeny. The phylogenetic tree was visualized in RStudio (version 4.4.2; RStudio, MA, USA) using ggtree (32).

### Statistical analysis

Descriptive statistics were used to summarize clinical characteristics, laboratory parameters, sequencing performance, and pathogen prevalence, reported as frequencies and percentages for categorical variables and medians with interquartile ranges (IQR) for continuous variables. The distribution of continuous variables was first assessed using the Shapiro–Wilk test to determine normality. Continuous variables were compared using the Wilcoxon rank-sum test. Microbial reads were processed after removing host sequences, and taxonomic relative abundances were normalized using total-sum scaling (TSS) to allow comparison across samples with variable sequencing depths. Alpha-diversity indices, including Shannon and Chao1, were calculated to assess within-sample microbial richness and diversity, while beta-diversity was evaluated using Bray–Curtis dissimilarity followed by principal coordinates analysis (PCoA). Differences in microbial community structure between clinical severity groups and other covariates were tested using PERMANOVA with 999 permutations. To identify risk factors associated with severe disease, logistic regression modeling was performed using the finalfit package in R. Univariable logistic regression analyses were initially performed, and variables with p < 0.20 were considered eligible for multivariable models. Statistical significance was defined as p < 0.05. All statistical analyses and visualizations were conducted using RStudio (version 4.4.2; RStudio, MA, USA).

## Ethics Approval

This study was conducted under ethical approvals obtained from the Research Ethics Committee of the Faculty of Associated Medical Sciences, Chiang Mai University (Approval No. AMSEC-66EM-011) and the Ethics Committee of the participating hospital (Approval No. 112/66). The research utilized de-identified clinical data and remnant respiratory specimens that were originally collected for routine diagnostic purposes. All samples were coded and anonymized prior to analysis to protect patient confidentiality. As only leftover diagnostic specimens were used and no interventions were conducted on patients, a waiver of informed consent was granted in accordance with Thai ethical guidelines for research use of archived specimens.

## Results

### Sequencing performance and pathogen profiles

To evaluate the performance of the optimized long-read mNGS workflow developed in our prior study, 101 respiratory samples from adult SARI patients were sequenced. Successful sequencing was achieved in 91% (92/101) of samples. Taxonomic classification at the family, genus, and species levels was obtained in 78% (79/101), 73% (74/101), and 72% (73/101) of samples, respectively (Figure 1A). Assembly failure occurred in 9% (9/101) of samples, while 13% (13/101) yielded host-derived reads only. To assess the added diagnostic value of mNGS, pathogen identification outcomes were compared with multiplex PCR results reported in our earlier investigation, as well as with conventional culture results from hospital records. The mNGS approach exhibited a notably higher detection rate (79/101; 78%) compared to multiplex PCR (47/101; 47%) and culture (40/101; 40%). Concordance analysis revealed that 30 samples (30%) were positive by both mNGS and multiplex PCR. Notably, mNGS uniquely identified pathogens in 18 samples (18%) that were negative by both multiplex PCR and culture (Figure 1B). To further explore the diagnostic utility of mNGS beyond targeted assays, we stratified the results based on multiplex PCR status. Among multiplex PCR-negative cases (n=34), mNGS detected a diverse array of clinically relevant pathogens, including *Klebsiella pneumoniae* (11.8%), *Haemophilus parainfluenzae* (8.8%), *Pseudomonas aeruginosa* (5.9%), SARS-CoV-2 (2.9%), and several oral anaerobes potentially implicated in lower respiratory tract infections in patients with comorbidities (Figure 1C). In multiplex PCR-positive samples (n=39), mNGS reliably confirmed the targeted pathogens while also uncovering additional co-infecting bacteria not included in the multiplex PCR panels (Figure 1D).

**Figure 1.**
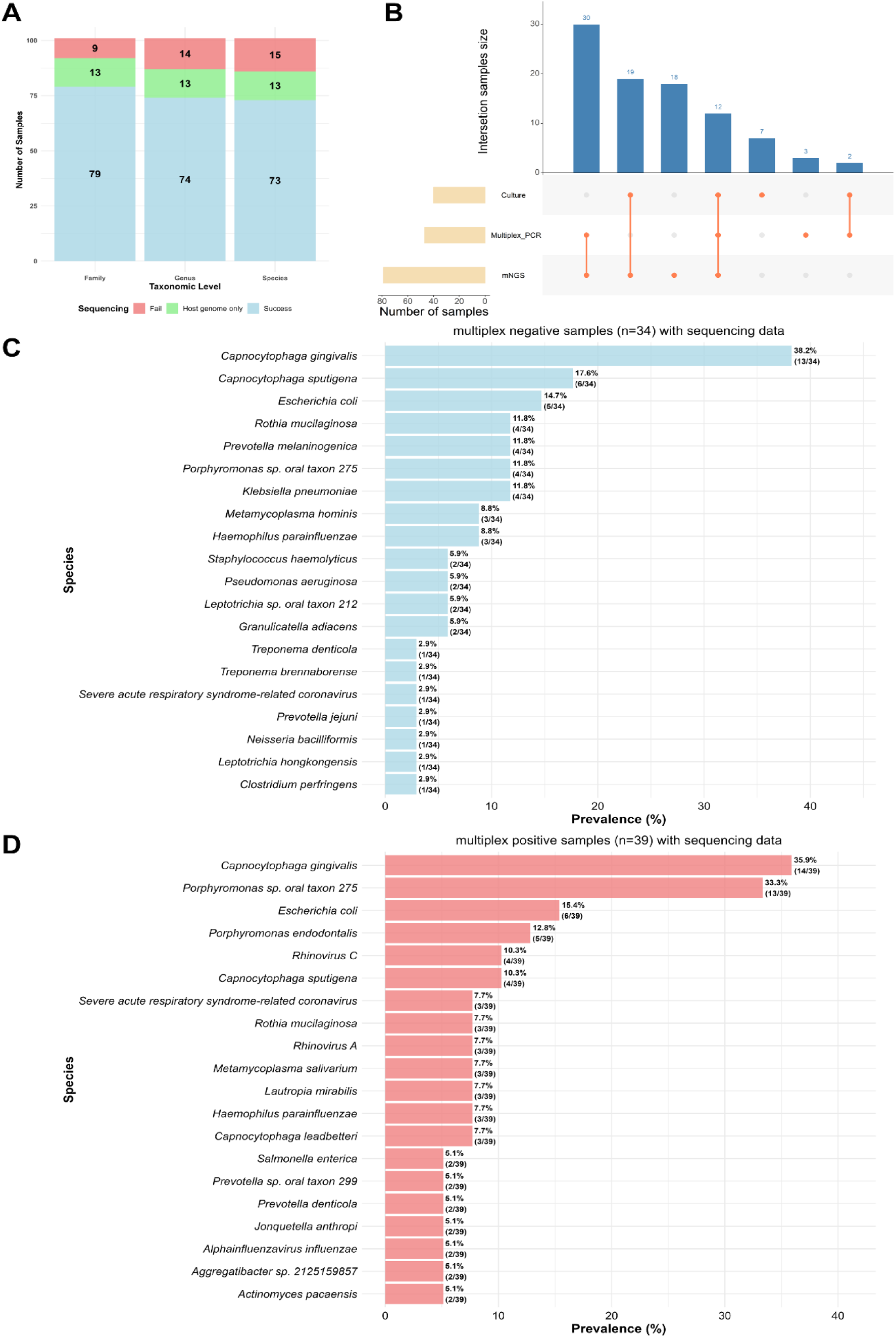
Summary of the optimized metagenomic sequencing performance, method concordance, and species-level taxonomic findings. **(A)** Sequencing outcomes for 101 samples at the family, genus, and species levels. Bar plot showing successful taxonomic assignment (blue), partial host-genome interference (green), and sequencing failure (red). **(B)** Upset plot showing pathogen detection across mNGS, multiplex PCR, and culture. The upper bars represent intersection sample sizes, while connected orange nodes indicate method combinations. Horizontal bars show the total number of positive samples per method. **(C)** Top 20 species identified by mNGS among multiplex PCR–negative samples (n=34). **(D)** Top 20 species identified by mNGS among multiplex PCR–positive samples (n=39).

### Microbial Landscape of SARI Patients

Metagenomic sequencing revealed a diverse and clinically significant microbial landscape among the 79 adult SARI patients (Figure 2). The most prevalent bacterial families included *Enterobacteriaceae* and *Flavobacteriaceae*, alongside several anaerobic and oral–respiratory–associated families such as *Porphyromonadaceae, Prevotellaceae, Neisseriaceae, and Pasteurellaceae*. Viral families were also represented, with *Picornaviridae* and *Coronaviridae* being the most commonly detected. At the genus level, *Capnocytophaga*, *Porphyromonas*, *Escherichia*, and *Prevotella* were predominant, collectively comprising a substantial portion of the detected microbial community. These genera reflect a blend of potential respiratory pathogens, oral anaerobes, and aspiration-associated taxa. Other notable genera included *Enterovirus*, *Rothia, Haemophilus, Klebsiella, Pseudomonas,* and *Leptotrichia*, all of which are recognized contributors to lower respiratory tract infections in adults. Species-level analysis further emphasized organisms of potential pathogenic importance. *Capnocytophaga gingivalis* was the most frequently detected species, followed by *Porphyromonas* sp. oral taxon 275, *Escherichia coli*, *Capnocytophaga sputigena*, *Rothia mucilaginosa*, and *Haemophilus parainfluenzae*. Several viral pathogens, including rhinovirus A, rhinovirus C, and SARS-CoV-2, were also identified. In addition, recognized bacterial respiratory pathogens such as *Klebsiella pneumoniae*, *Prevotella melaninogenica*, and *Metamycoplasma hominis* were detected, highlighting the complex and polymicrobial nature of SARI infections in this cohort.

**Figure 2.**
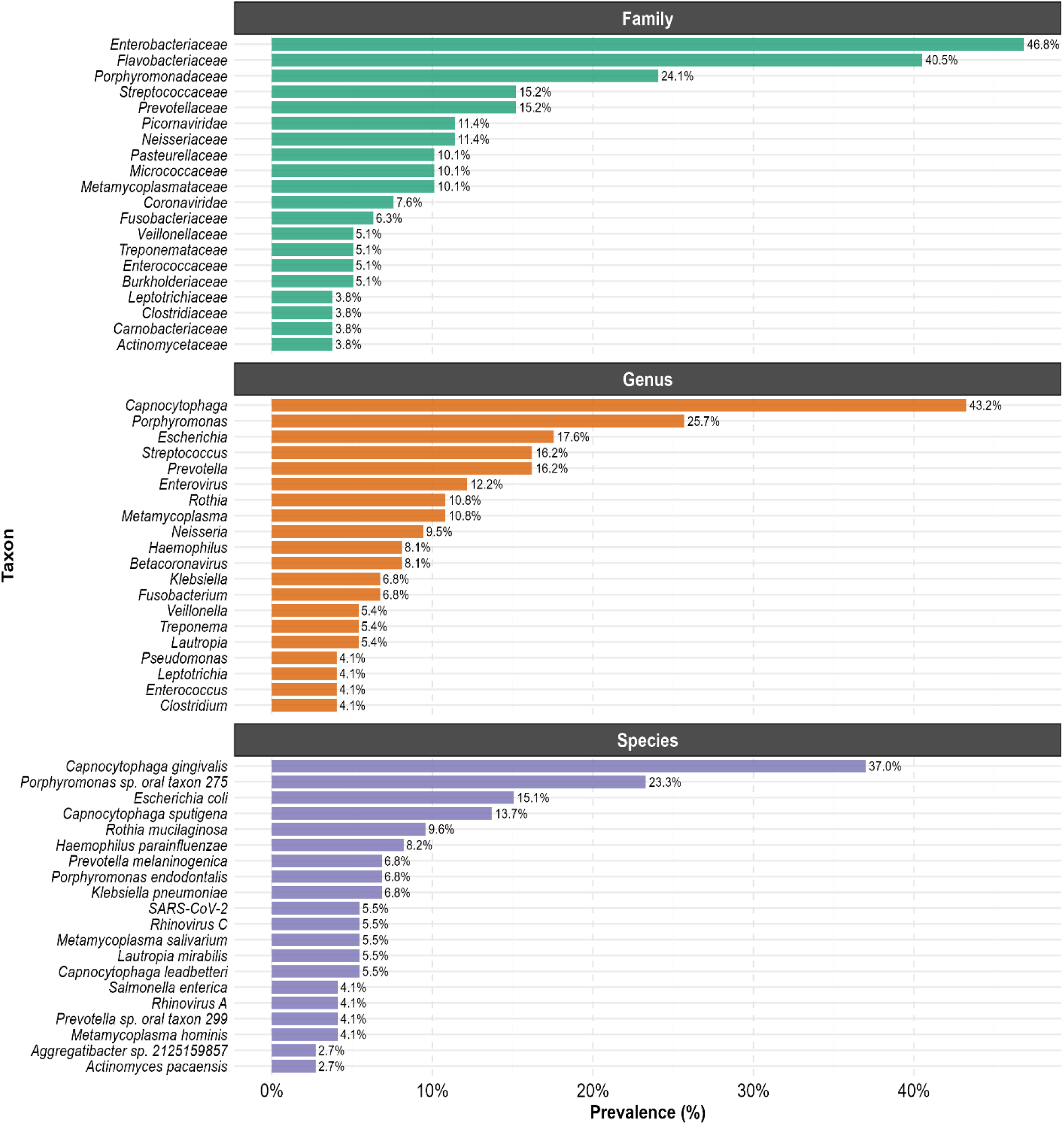
Prevalence of identified taxa among adult SARI patients based on metagenomic sequencing. Horizontal bar plots illustrate the distribution of detected pathogens at three taxonomic levels: family, genus, and species. Only the top 20 taxa at each level are displayed. Prevalence of each taxon was calculated as the proportion of samples with successful mNGS classification (n=79). Samples with host-only reads (n=13) or assembly failure (n=9) were excluded from denominator calculations.

### Comparison of pathogen detection across mNGS, multiplex PCR, and culture methods

To assess the concordance between standard diagnostic methods (multiplex PCR and culture) and the optimized metagenomic sequencing approach, we compared species-level pathogen identifications across the three platforms (Figure 3). The mNGS approach demonstrated the broadest detection range, identifying a substantially more diverse array of viral and bacterial taxa compared to culture or multiplex PCR. Multiplex PCR showed strong performance for viral agents included in its detection panel but identified fewer bacterial species compared to mNGS and culture. Culture-based methods predominantly recovered fast-growing respiratory bacteria. Overlapping detections were mainly observed between mNGS and multiplex PCR for high-abundance viral species such as rhinovirus, human metapneumovirus, influenza virus, human coronaviruses, and human parainfluenza virus, demonstrating strong agreement for high-abundance respiratory viruses. In contrast, concordance between mNGS and culture was confined to culturable organisms, particularly *Klebsiella pneumoniae* and *Acinetobacter baumannii*, though these were detected in relatively few samples. Importantly, mNGS identified atypical pathogens such as *Mycoplasma pneumoniae*, which are typically challenging to culture and require molecular assays for confirmation. Furthermore, several pathogens were uniquely detected by mNGS, highlighting its capability to uncover unculturable, rare, or unexpected taxa beyond the scope of conventional targeted diagnostics.

**Figure 3.**
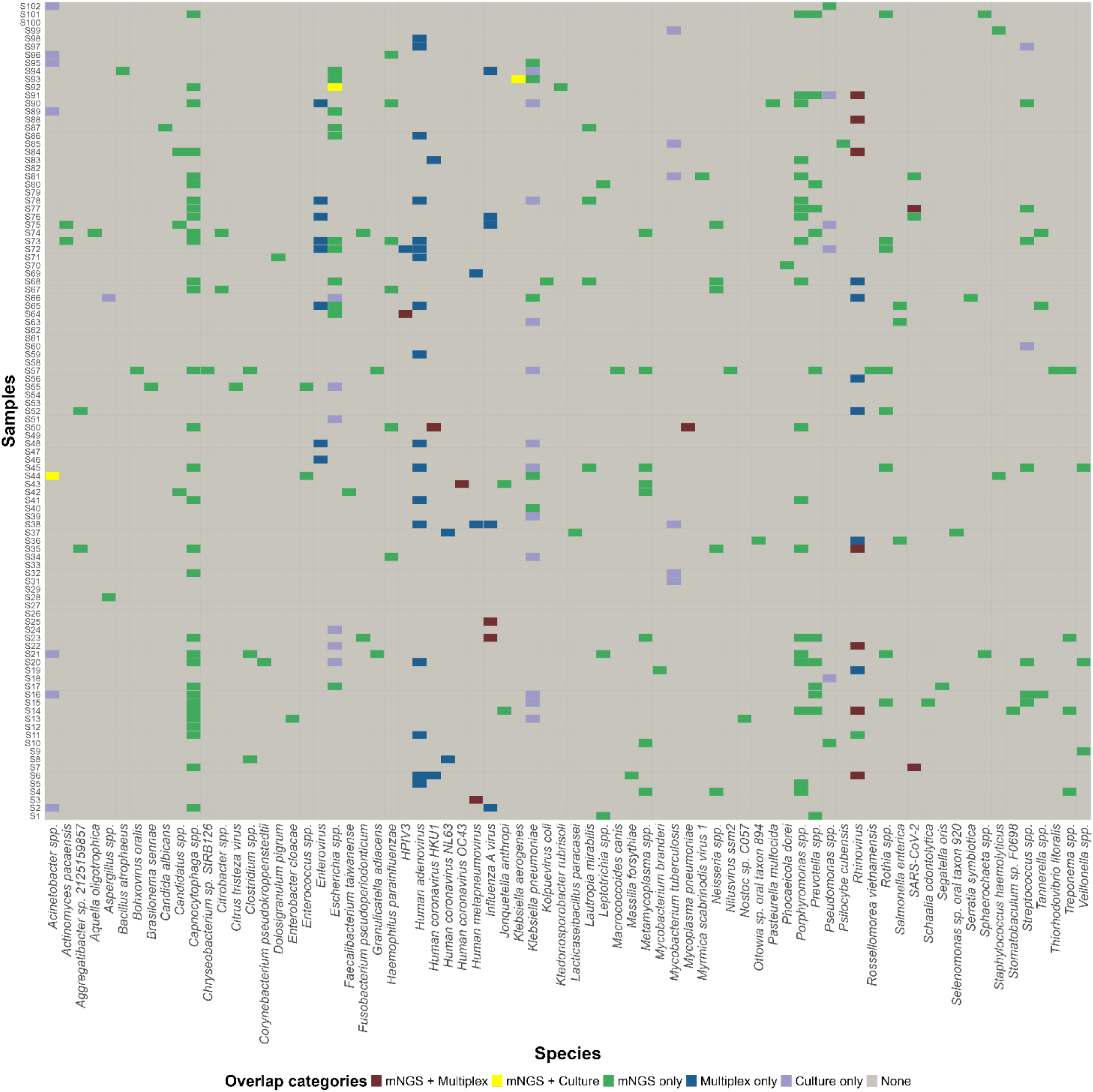
Comparison of pathogen detection across three diagnostic methods. Tile plot comparing organisms detected using mNGS, multiplex PCR, and culture among SARI patient samples. Rows represent patient samples, and columns represent detected species. Colors indicate species detected by mNGS + multiplex PCR (red), mNGS + culture (yellow), mNGS only (green), multiplex only (blue), culture only (purple), or no detection (grey). mNGS detected the highest diversity of pathogens, including atypical bacteria and unculturable pathogens, while multiplex PCR predominantly identified targeted viral species within the panel, and culture recovered only a limited subset of classical bacterial pathogens.

### Genome-wide coverage profiles confirm accurate pathogen detection by mNGS

To evaluate the reliability and accuracy of pathogen detection achieved by the optimized mNGS workflow, genome-wide coverage profiles were generated for representative pathogens with concordant detection by both multiplex PCR and mNGS. These analyses demonstrated robust and contiguous read coverage across the genomes of various respiratory pathogens, including *Mycoplasma pneumoniae*, human coronaviruses (HKU1, OC43), human metapneumovirus, human parainfluenza virus 3, human rhinovirus, influenza A virus, and SARS-CoV-2 (Figure 4). High-density, continuous coverage, particularly for rhinovirus and influenza A, confirmed effective enrichment and recovery of pathogen reads, even in the context of sputum samples characterized by high human genomic background. For lower-abundance pathogens such as *M. pneumoniae* and coronaviruses (HKU1, OC43), the read coverage patterns remained specific and aligned precisely to their respective reference genomes, supporting confident and accurate identification.

**Figure 4.**
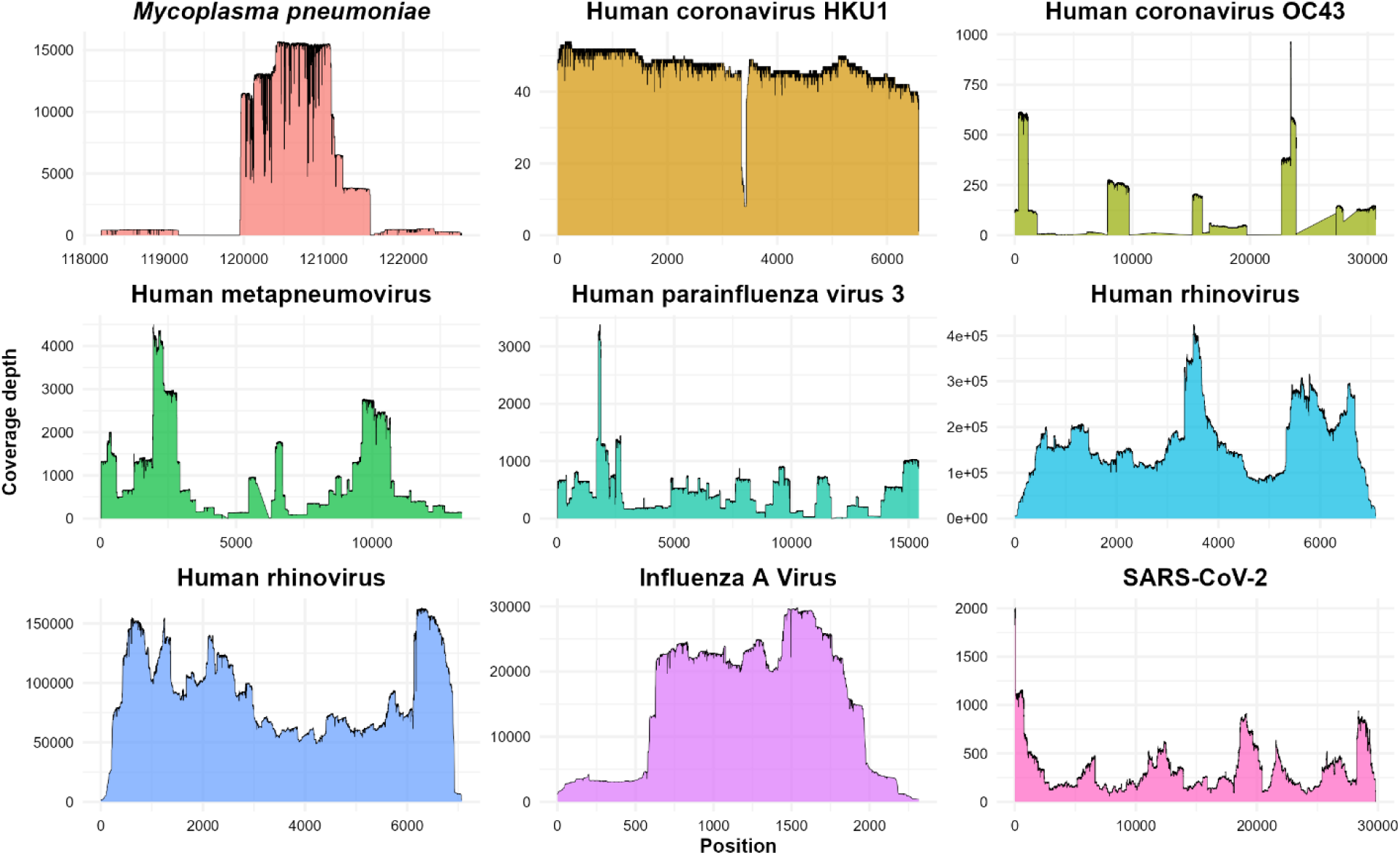
Metagenomic coverage depth across representative respiratory pathogen genomes. Coverage plots show read depth across reference genomes for pathogens identified in representative SARI patient samples. Reads were mapped to respective reference genomes using minimap2, and per-base depth was visualized after host reads removal. Distinct coverage patterns are observed for each pathogen, with high and continuous depth for viruses such as rhinovirus and influenza A and characteristic localized patterns for *M. pneumoniae* and coronaviruses.

### Age-stratified pathogen profile

To investigate age-specific distributions of respiratory pathogens among adult SARI patients and assess whether certain pathogens disproportionately affect specific age groups, we conducted an age-stratified analysis of metagenomic findings (Figure 5). The results revealed notable variations in pathogen prevalence across the three defined age groups: 18–30 years, 31–64 years, and ≥65 years. Overall, elderly patients (≥65 years) exhibited the greatest diversity of detected pathogens, suggesting a more complex microbial landscape in this population. Several organisms, including *Haemophilus parainfluenzae*, *Klebsiella pneumoniae*, and rhinovirus, were consistently detected across all age groups, indicating their potential role as common etiologic agents in adult SARI. In younger adults (18–30 years), pathogens such as human coronavirus HKU1, human metapneumovirus, and *Mycoplasma pneumoniae* were more frequently identified. Conversely, *Acinetobacter baumannii* was predominantly found in the elderly cohort, likely reflecting higher rates of healthcare-associated exposures in this age group. In the intermediate age group (31–64 years), increased frequencies of human coronavirus OC43 and *Klebsiella aerogenes* were observed. Despite these trends, no single pathogen was uniformly dominant across all age groups, highlighting the heterogeneous etiology of SARI in adults and the influence of host factors such as age and comorbidity on pathogen distribution.

**Figure 5.**
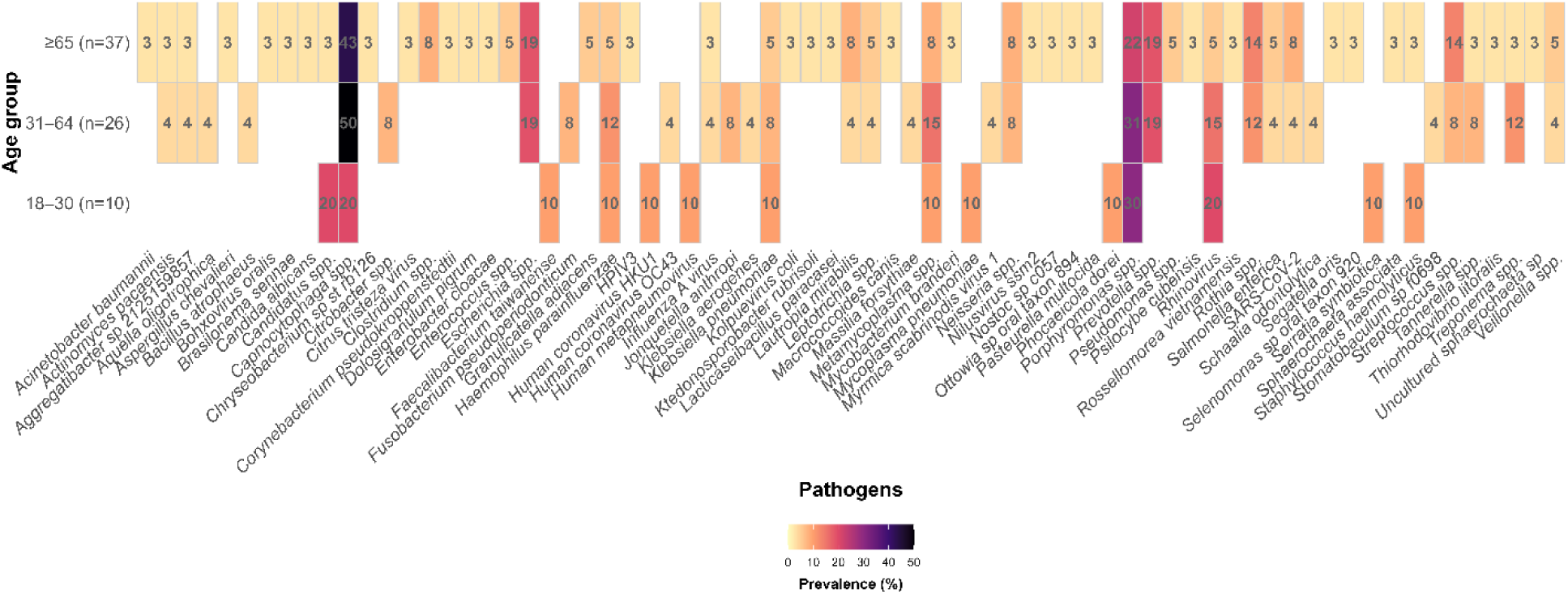
Age-stratified prevalence of respiratory pathogens detected by metagenomic sequencing in SARI patients. Heatmap showing the prevalence (%) of each detected pathogen across three age groups: 18–30 years (n=10), 31–64 years (n=26), and ≥65 years (n=37). The color gradient represents pathogen prevalence from low (light yellow) to high (dark purple), and numeric values indicate the percentage of positive samples within each age group for that pathogen. Pathogens are grouped at the species level and shown along the x-axis.

### Laboratory markers by disease severity

To assess whether hematological parameters could differentiate disease severity in SARI patients, we compared white blood cell (WBC), lymphocyte, neutrophil, and platelet counts between mild and severe cases (Figure 6). Severe cases exhibited marked lymphopenia (p < 0.01), elevated neutrophil counts (p < 0.05), and significantly increased total WBC counts (p < 0.01), consistent with acute inflammatory responses and potential secondary bacterial infection. In contrast, platelet counts did not significantly differ between the severity groups, indicating that thrombocytopenia was not a distinguishing feature in this cohort. These hematological patterns—characterized by lymphopenia, neutrophilia, and leukocytosis—may serve as useful clinical indicators of severe disease and could support early risk stratification and management decisions in adult SARI patients.

**Figure 6.**
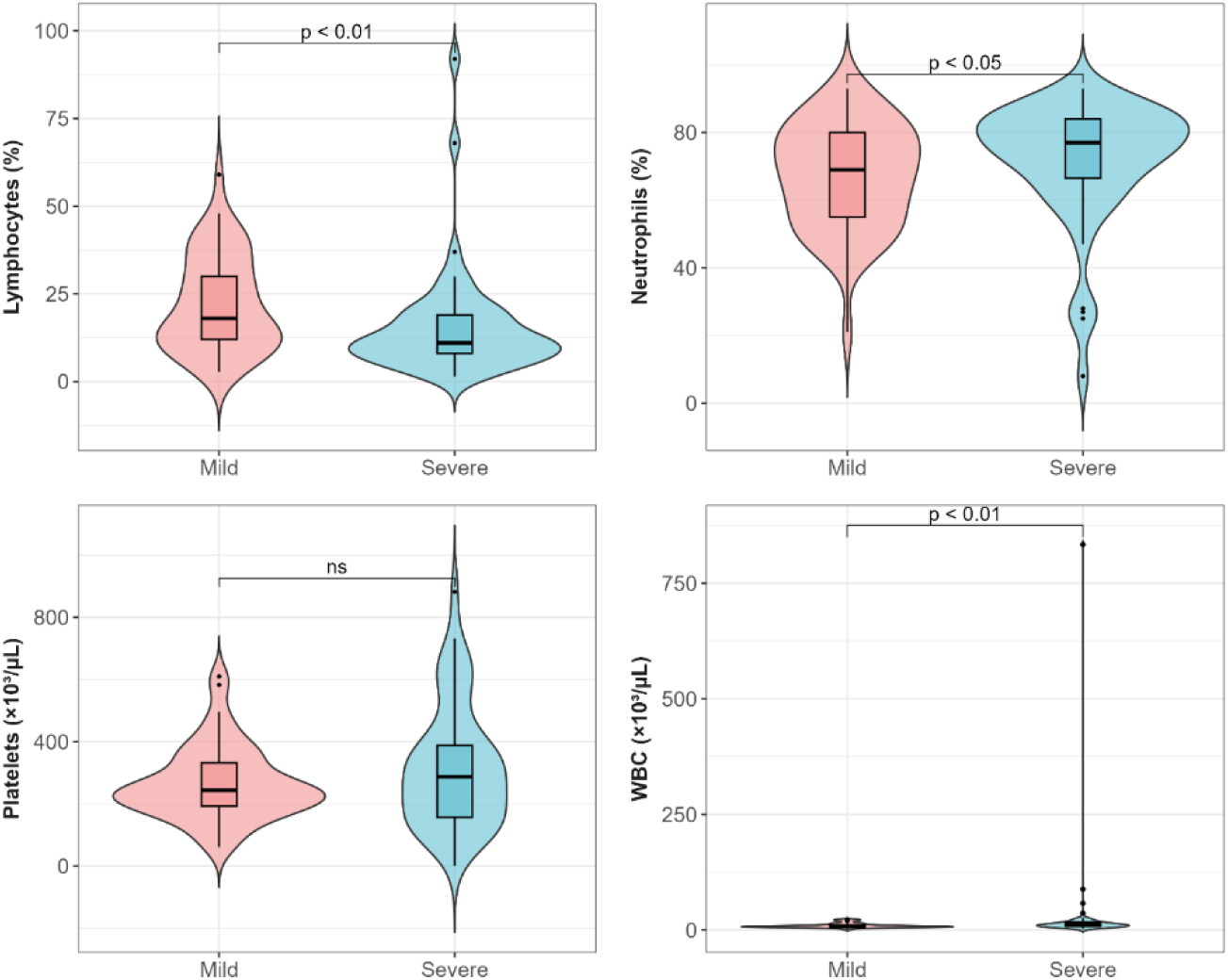
Hematological profiles of SARI patients by disease severity. Violin plots show distributions of **(A)** lymphocyte, **(B)** neutrophil, **(C)** platelets, and **(D)** white blood cell count among mild and severe SARI cases. Overlaid boxplots represent median values and interquartile ranges. Statistical comparisons were performed using the Wilcoxon rank-sum test due to non-normal distributions. Significant differences were observed for lymphocytes (p < 0.01), neutrophils (p < 0.05), and WBC count (p < 0.01), while platelets showed no significant difference. WBC, white blood cell; ns, no significant.

### Microbial diversity analysis

To determine whether microbial community diversity correlates with clinical severity and may serve as an indicator of disease progression, we analyzed alpha and beta diversity among mild and severe SARI cases. Alpha diversity metrics, including species richness and evenness, were assessed using the Chao1 and Shannon indices. For the mild group, the median Chao1 index was 3 (IQR: 2–7), and the Shannon index was 0.45 (IQR: 0.14–0.88). In the severe group, the corresponding values were 3 (IQR: 2–6) and 0.45 (IQR: 0.22–0.90), respectively. No statistically significant differences were observed between the groups for either index (Chao1: p = 0.85; Shannon: p = 0.49) (Figure 7A). Beta diversity was evaluated using Bray–Curtis distances, with principal coordinates analysis (PCoA) illustrating the between-sample variation in microbial composition (Figure 7B). The ordination plot showed no distinct clustering by severity category, indicating similar microbial community structures across groups. This was further supported by PERMANOVA analysis, which yielded a non-significant result (R² = 0.013, p = 0.142). These results suggest that disease severity in this adult SARI cohort is not driven by broad shifts in microbial diversity. Instead, severity may be more closely associated with the presence of specific pathogenic taxa or host factors.

**Figure 7.**
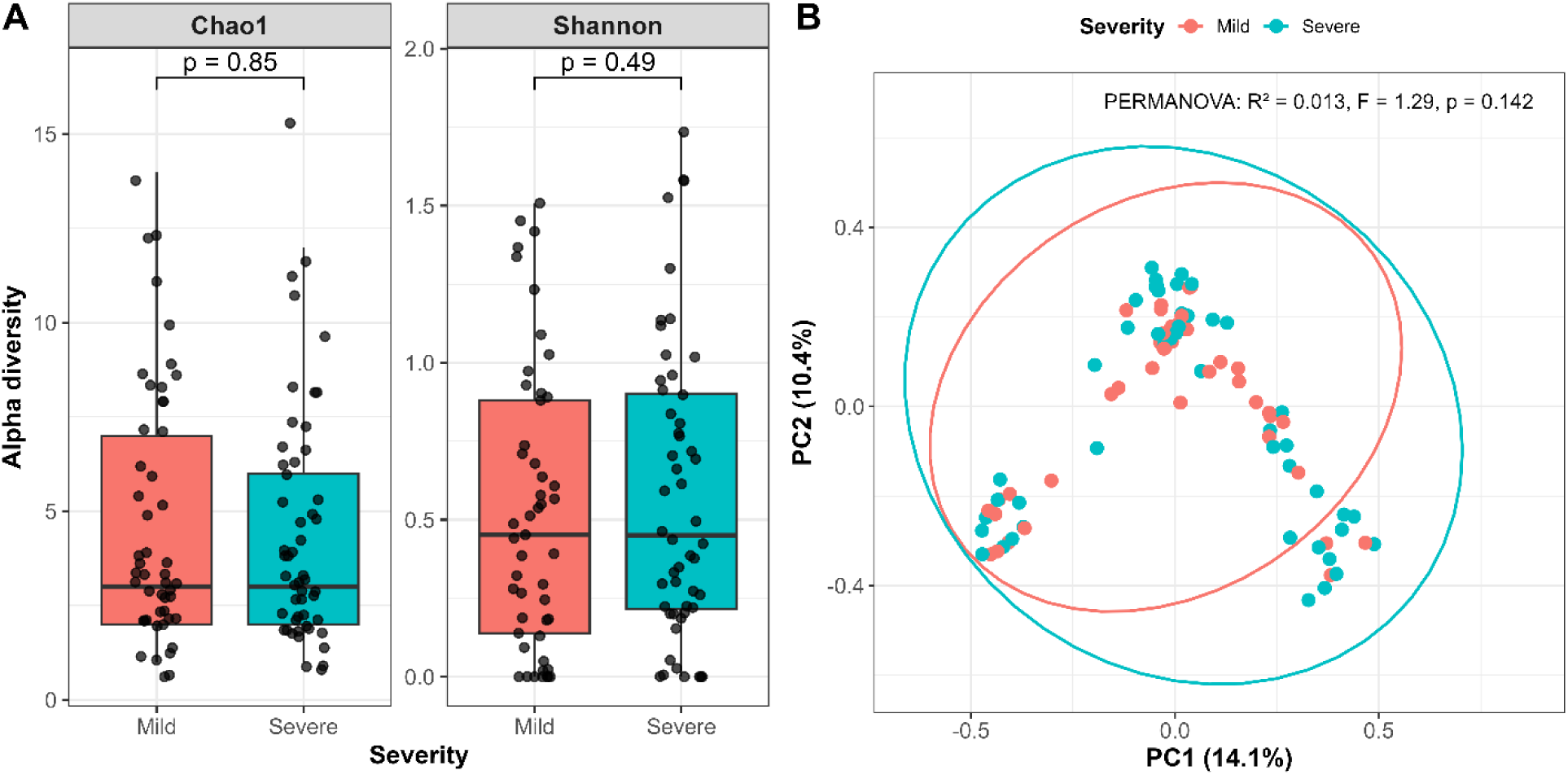
Alpha and beta diversity of respiratory microbial communities stratified by disease severity. **(A)** Boxplots showing alpha diversity indices (Chao1 richness and Shannon diversity) for mild and severe SARI cases. Horizontal lines within the boxplots indicate median values, and p-values were obtained using the Wilcoxon rank-sum test. **(B)** Principal Coordinates Analysis (PCoA) plot based on Bray–Curtis dissimilarity illustrating beta diversity between mild and severe groups. Ellipses represent 95% confidence regions for each group. Statistical significance was assessed using PERMANOVA (R² = 0.013, F = 1.29, p = 0.142).

### Pathogen-Specific Associations with Outcomes

To explore whether specific microbial signatures correlate with disease severity, we conducted a heatmap-based analysis of species-level relative abundance across fungal, bacterial, and viral taxa (Figure 8). The results revealed substantial inter-patient variability, with diverse viral and bacterial profiles observed throughout the cohort. Viral pathogens, including human metapneumovirus, human parainfluenza virus, human coronaviruses, influenza A virus, and rhinovirus, were commonly detected in patients with mild diseases. In contrast, individuals with pneumonia or those requiring special care frequently exhibited enrichment of nosocomial or opportunistic bacterial pathogens such as *Klebsiella pneumoniae*, *Haemophilus parainfluenzae*, *Klebsiella aerogenes*, *Acinetobacter baumannii*, and *Staphylococcus haemolyticus*, suggesting possible bacterial mono- or super-infections contributing to increased disease severity. SARS-CoV-2 was identified in two fatal cases, and *Klebsiella pneumoniae* was detected in one of these, underscoring their potential role in adverse outcomes. Fungal species were broadly distributed across cases, although *Candida spp.* was more abundant in severe cases. However, fungal presence did not demonstrate a consistent outcome-specific enrichment pattern.

**Figure 8.**
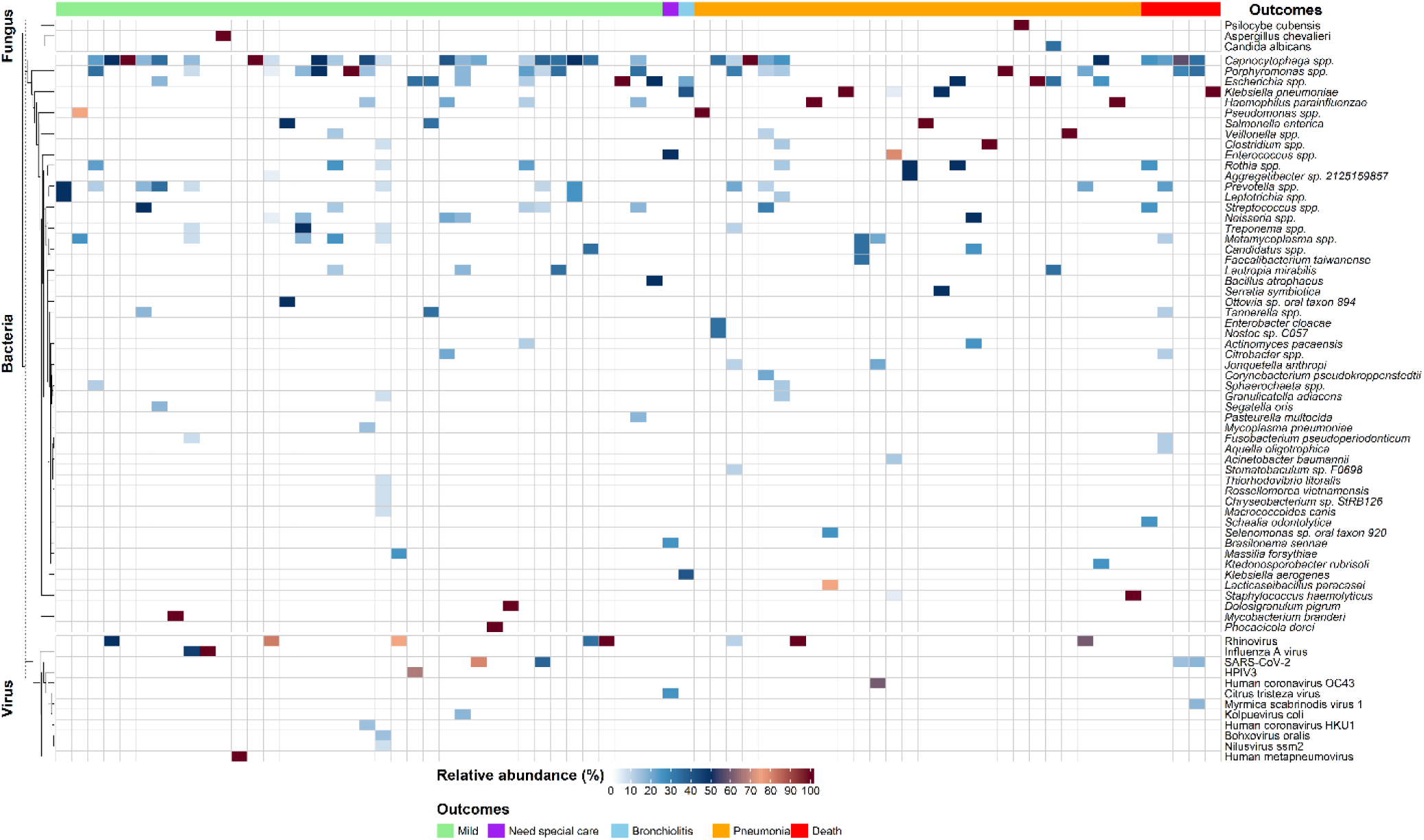
Distribution and relative abundance of respiratory pathogens across clinical outcomes in adult SARI cases. Heatmap showing the relative abundance of fungal, bacterial, and viral taxa across SARI patient samples. Columns represent individual patient samples, annotated by clinical outcomes (mild, need special care, bronchiolitis, pneumonia, and death). Each row represents the identified species. Microbial taxa are grouped into fungal, bacterial, and viral domains and arranged based on hierarchical clustering. Color gradients indicate relative abundance per sample after host-read removal and normalization.

### Risk factors associated with disease severity in SARI patients

To identify risk factors associated with severe outcomes among SARI patients, we conducted both univariable and multivariable logistic regression analysis incorporating demographic features, comorbidities, vaccination status, and clinical parameters (Supplementary Table 1). Male sex was significantly associated with increased risk of severe disease compared to female sex (odds ratio (OR) = 3.56, 95% confidence interval (CI): 1.09–13.21, p = 0.043), suggesting possible underlying immunological or behavioral factors influencing susceptibility. In contrast, higher oxygen saturation levels were strongly protective, with each unit increase associated with reduced odds of severe illness (OR = 0.54 per unit increase, 95% CI: 0.39–0.71, p < 0.001) (Figure 9). Notably, influenza vaccination status was not significantly associated with reduced severity in this cohort. Other clinical factors, including pre-existing pulmonary disease, hypertension, and overall comorbidity burden, also did not demonstrate statistically significant associations with severe disease.

**Figure 9.**
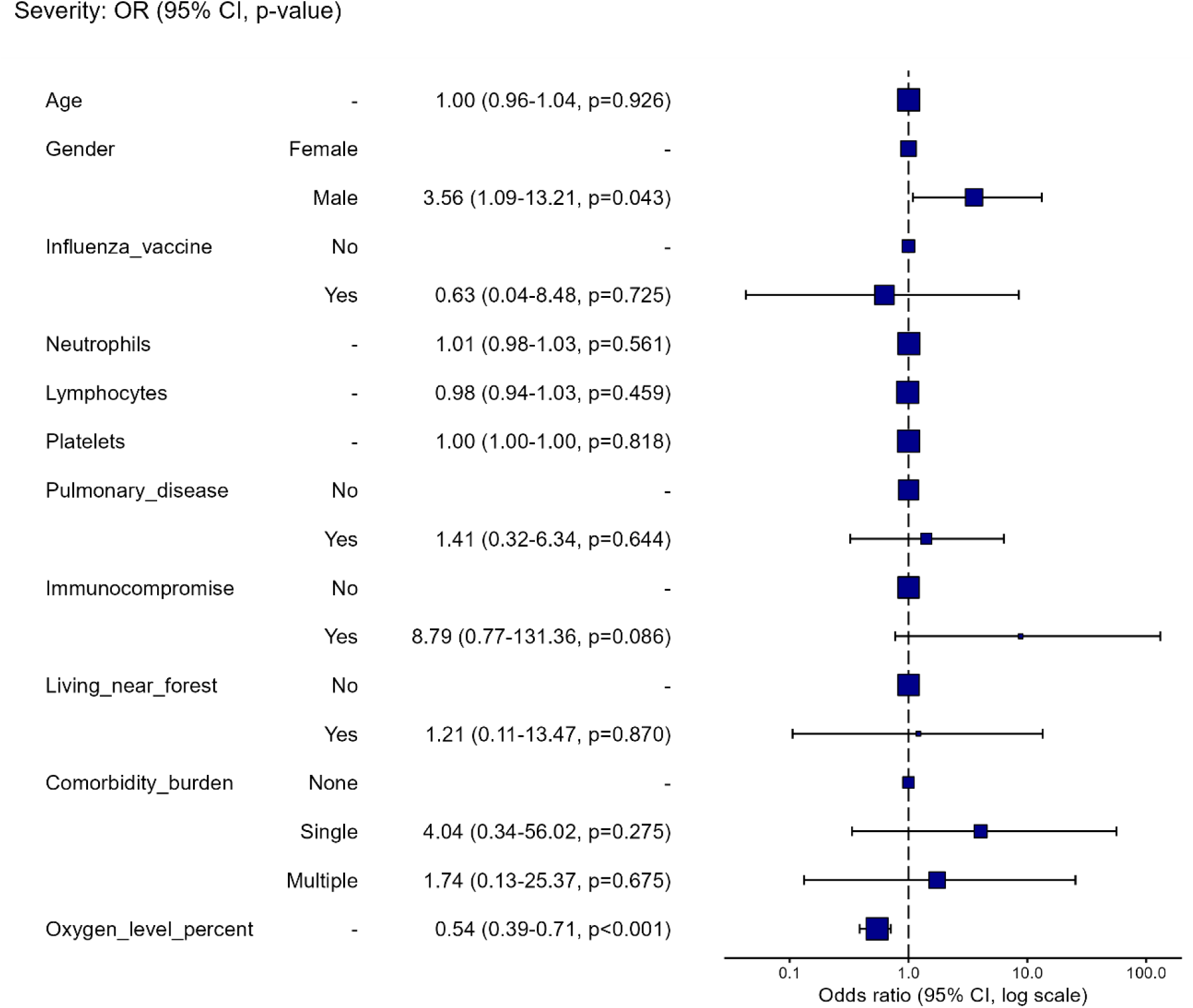
Multivariable logistic regression analysis for factors associated with severe outcomes among SARI patients in Northern Thailand. The forest plot illustrates odds ratios (OR) with 95% confidence intervals on a logarithmic scale. Variables include patient demographics, vaccination status, comorbidities, and clinical parameters. A vertical dashed line represents an OR of 1.00, indicating no association. Increased risk of disease severity was observed among male patients, whereas higher oxygen saturation was strongly associated with reduced disease severity.

### Phylogenetic characterization of rhinovirus

To characterize the circulating human rhinovirus (HRV) strains associated with adult SARI during the 2023–2024 period, we conducted phylogenetic analysis on all HRV-positive samples (n=8). The resulting maximum-likelihood tree revealed distinct clustering of HRV strains into three species: HRV-A (n=3), HRV-B (n=1), and HRV-C (n=4), as illustrated in Figure 10. Among the HRV-C strains, 4 clinical samples were grouped into genotypes C22 and C42. The 3 HRV-A sequences clustered within the HRV-A1 subgroup, aligning with the closest reference sequences. One sample mapped to the HRV-B lineage and showed close phylogenetic proximity to HRV-B48. Enterovirus D68 (EV-D68) was included as an evolutionary outgroup.

**Figure 10.**
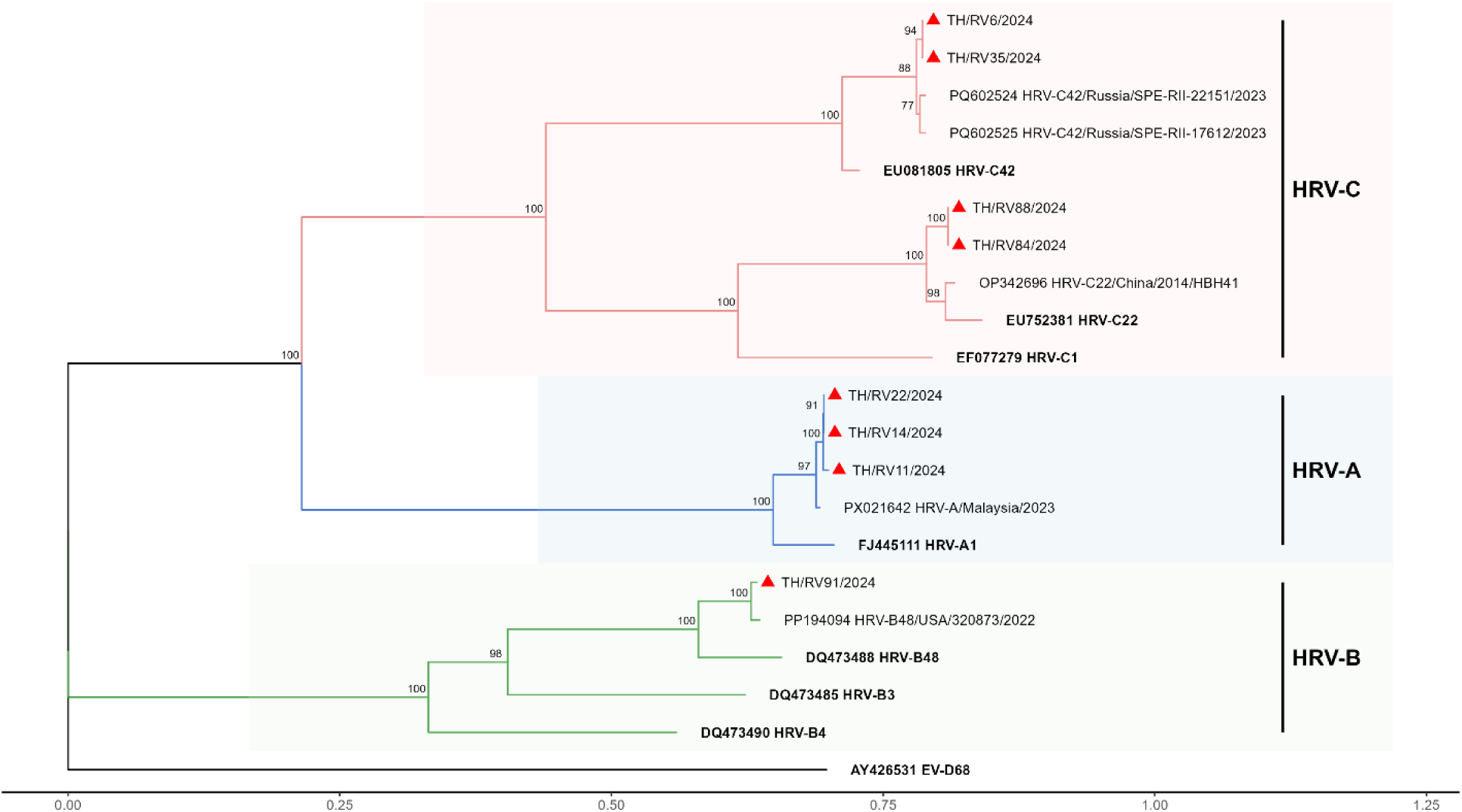
Phylogenetic analysis of rhinovirus-positive samples among SARI patients. A maximum-likelihood tree demonstrates distinct clustering of HRV-A (blue), HRV-B (green), and HRV-C (red) clades. Samples from this study (marked with red triangles) were grouped with established prototype strains, including HRV-C42, HRV-C22, HRV-A1, and HRV-B48. Reference genomes are shown in bold letters. Enterovirus D68 (EV-D68) was included as an outgroup. Bootstrap values are displayed along branches.

## Discussion

This study highlights the substantial clinical and diagnostic utility of long-read metagenomic next-generation sequencing (mNGS) for comprehensive pathogen detection in adult patients hospitalized with severe acute respiratory infections (SARI) in northern Thailand. An optimized nanopore-based sequencing workflow, applied to sputum specimens, achieved a diagnostic yield of 78%, significantly outperforming multiplex PCR (47%) and culture (40%). These results underscore the advantages of unbiased sequencing approaches in identifying both expected and atypical pathogens, especially in settings where conventional diagnostics are limited by targeted panels or low sensitivity.

Notably, mNGS identified additional pathogens in 18% of cases that were negative by both PCR and culture, including clinically relevant organisms such as *Klebsiella pneumoniae, Haemophilus parainfluenzae, Pseudomonas aeruginosa*, and even SARS-CoV-2. These findings emphasize the limitations of targeted panels, which cannot detect pathogens outside of their scope, and support the role of mNGS in uncovering unanticipated etiologies. The detection of SARS-CoV-2 in PCR-negative samples may reflect genomic variation affecting primer-binding regions, highlighting the need for continuous molecular surveillance (33, 34). Conversely, certain pathogens detected by culture or PCR were missed by mNGS, likely due to low microbial biomass, high host background, or degraded nucleic acids (35, 36).

Concordance between mNGS and multiplex PCR was particularly strong for common respiratory viruses, and genome-wide coverage profiles confirmed high-confidence detection, including complete or near-complete coverage for *rhinovirus*, *human metapneumovirus*, and *coronaviruses*. Even low-abundance targets such as *Mycoplasma pneumoniae* exhibited specific, although discontinuous, coverage patterns consistent with true positives. *M. pneumoniae* is difficult to culture and often requires molecular tests such as PCR assays for diagnosis (37). The ability of mNGS to detect this atypical pathogen without prior targeting is useful, particularly for community-acquired atypical pneumonia (38).

Moreover, polymicrobial infections were frequently detected in SARI patients, consisting of opportunistic, nosocomial, and oral anaerobes, with the predominance of families such as *Enterobacteriaceae*, *Flavobacteriaceae*, *Porphyromonadaceae*, and *Prevotellaceae*. Although some may indicate oropharyngeal colonization, their predominance in severe cases, especially among elderly or comorbid patients, suggests the potential for aspiration-associated pneumonia or secondary bacterial superinfection (39, 40).

Age-stratified analysis revealed distinct pathogen patterns across age groups. Younger adults exhibited elevated detection rates of HMPV, HCoV-HKU1, and Mycoplasma pneumoniae, pathogens commonly associated with community-acquired respiratory infections (41, 42). Middle-aged adults showed moderate frequency of HCoV-OC43 and *Klebsiella aerogenes*, while the elderly exhibited a broader pathogen diversity, with increased detection of *Acinetobacter baumannii*, a healthcare-associated infection (43). The identification of *Haemophilus parainfluenzae* and *Klebsiella pneumoniae* in all age groups indicates that they are ubiquitous opportunistic pathogens in SARI. These patterns indicate varying exposures, immune responses, and comorbidity burdens among different age groups, which may influence respiratory microbial colonization and infection risk.

Among viral etiologies, rhinovirus emerged as a predominant pathogen within this cohort. Phylogenetic analysis indicated the circulation of HRV-A (37.5%), HRV-B (12.5%), and HRV-C (50%), with HRV-C being the most common. This is consistent with a recent study in Thailand, where 45% of HRV-C is found in SARI patients (44). Recent studies also implicate HRV-C in severe lower respiratory tract infections, particularly in adults and children with chronic lung disease or immunosuppression (45, 46). However, the molecular epidemiology of rhinovirus in SARI warrants investigation, as certain clades (e.g., HRV-C) may increase virulence for the lower airways. These results highlight the potential of mNGS to facilitate real-time molecular surveillance of circulating HRV strains in Thailand.

Despite detailed microbial profiling, alpha and beta diversity analyses showed no significant differences between mild and severe cases, suggesting that overall microbial diversity is not a major determinant of clinical severity in adult SARI. Instead, disease severity may be more influenced by specific pathogenic taxa, underlying host factors, and immunological responses rather than by broad alterations in community ecology alone. In severe and fatal cases, specific pathogenic taxa, such as *Klebsiella pneumoniae, Acinetobacter baumannii, Staphylococcus haemolyticus,* and *Klebsiella aerogenes*, were commonly found. These patterns align with viral–bacterial synergy, wherein viral infection facilitates subsequent bacterial invasion (47, 48). Notably, SARS-CoV-2 was detected in two fatal cases and *Klebsiella pneumoniae* in one, emphasizing their pathogenic significance in SARI mortality.

Multivariable logistic regression identified male sex and decreased oxygen saturation as independent predictors of severe disease. The association between being male and having worse outcomes may be due to differences in the immune system, hormones, or other behavioral factors (49, 50). Hypoxemia indicates impaired gas exchange and thus serves as a readily measurable clinical biomarker for risk stratification and determinations related to ICU admission or mechanical ventilation (51).

Overall, this study supports the integration of long-read mNGS into respiratory diagnostics and public health surveillance. It facilitates the detection of a wide array of pathogens—including unculturable or unanticipated organisms—and enables real-time monitoring of pathogen evolution and transmission. This capability is particularly valuable for pandemic preparedness and outbreak response. Nevertheless, some limitations still remain in this study. First, the single-center setting may limit generalizability, particularly given regional variations in pathogen epidemiology, antimicrobial resistance patterns, and healthcare practices. Multicenter validation studies encompassing diverse geographic and demographic populations are warranted. Second, mNGS cannot independently differentiate colonization and true infection, particularly for organisms commonly found in the upper respiratory tract or oral microbiome; thus, integration with clinical data remains essential. Third, the absence of a healthy control group hindered the evaluation of background microbial colonization patterns, which would have enhanced the inference regarding pathogen-specific associations with SARI. Future studies involving healthy controls will be crucial for establishing baseline respiratory microbial signatures. Finally, while mNGS detected a broad array of organisms, functional metagenomic or metatranscriptomic approaches would provide deeper insights into microbial gene expression, virulence factor activity, and metabolic interactions within the respiratory microbiome. Future research focusing on investigation of host-pathogen interactions through integrative multi-omics approaches combining metagenomics, transcriptomics, and immunoprofiling would provide deeper insight into disease mechanisms.

## Conclusions

This study demonstrates that long-read metagenomic sequencing is a robust and comprehensive diagnostic platform for the etiological investigation of severe acute respiratory infections (SARI) in adults. By enabling the detection of a broad spectrum of viral, bacterial, and atypical pathogens, including those frequently missed by conventional methods, mNGS significantly enhances diagnostic yield and clinical insight. Its capacity to identify co-infections and provide strain-level resolution further underscores its value in both clinical diagnostics and infectious disease surveillance. Importantly, the severity of SARI was more closely associated with specific pathogens and host-related factors than with overall microbial diversity, emphasizing the need for pathogen-specific and host-aware clinical management strategies. These findings support the integration of long-read mNGS into respiratory infection diagnostics and real-time pathogen surveillance frameworks. Future research incorporating healthy controls, multi-omics approaches, and multicenter cohorts will be critical to refine the clinical interpretation of mNGS data and to advance its application in outbreak preparedness and precision infectious disease management.

## Data Availability

All data produced in the present study are available upon reasonable request to the authors

## Acknowledgements

We gratefully acknowledge Nakornping hospital for providing clinical specimens, and the LUCENT international collaboration, Associated Medical Sciences, Chiang Mai University, for providing laboratory facilities and technical support. N.K.K. was a candidate in PhD Program in Biomedical Sciences, Faculty of Associated Medical Sciences, Chiang Mai University, under the CMU Presidential Scholarship.

