## Supplementary data for "Comprehensive Pathogen Profiling in Adult Patients with Severe Acute Respiratory Infections Using Metagenomic Next-Generation Sequencing of Sputum Samples"

**Supplementary Table 1.** Factors associated with severe outcomes in patients with severe acute respiratory infection.

| **Dependent: Severity** |  | **Mild** | **Severe** | **OR (univariable)** | **OR (multivariable)** |
| --- | --- | --- | --- | --- | --- |
| Age | Mean (SD) | 54.0 (21.7) | 61.4 (18.1) | 1.02 (1.00-1.04, p=0.068) | 1.00 (0.96-1.04, p=0.926) |
| Gender | Female | 24 (54.5) | 20 (45.5) | - | - |
|  | Male | 25 (43.9) | 32 (56.1) | 1.54 (0.70-3.42, p=0.29) | 3.56 (1.09-13.21, p=0.043) |
| Occupation | No occupation | 15 (42.9) | 20 (57.1) | - | - |
|  | Farmer | 1 (33.3) | 2 (66.7) | 1.50 (0.13-34.07, p=0.75) | - |
|  | employee | 4 (80.0) | 1 (20.0) | 0.19 (0.01-1.43, p=0.20) | - |
|  | State enterprises employee | 2 (40.0) | 3 (60.0) | 1.13 (0.17-9.36, p=0.90) | - |
|  | Civil servant | 5 (50.0) | 5 (50.0) | 0.75 (0.18-3.15, p=0.69) | - |
|  | Work for wages | 10 (40.0) | 15 (60.0) | 1.12 (0.40-3.24, p=0.83) | - |
|  | Merchant | 5 (62.5) | 3 (37.5) | 0.45 (0.08-2.13, p=0.32) | - |
|  | Student | 6 (66.7) | 3 (33.3) | 0.37 (0.07-1.66, p=0.21) | - |
| Influenza vaccine | No | 17 (63.0) | 10 (37.0) | - | - |
|  | Yes | 32 (43.2) | 42 (56.8) | 2.23 (0.91-5.68, p=0.083) | 0.63 (0.04-8.48, p=0.725) |
| Oxygen level percent | Mean (SD) | 94.1 (1.9) | 88.6 (6.8) | 0.58 (0.45-0.72, p<0.001) | 0.54 (0.39-0.71, p<0.001) |
| WBC counts | Mean (SD) | 9108.2 (4961.0) | 30635.3 (115585.3) | 1.00 (1.00-1.00, p=0.015) | - |
| Neutrophils | Mean (SD) | 61.4 (25.4) | 68.5 (24.4) | 1.01 (1.00-1.03, p=0.158) | 1.01 (0.98-1.03, p=0.561) |
| Lymphocytes | Mean (SD) | 19.2 (14.5) | 13.9 (14.0) | 0.97 (0.94-1.00, p=0.077) | 0.98 (0.94-1.03, p=0.459) |
| Platelets | Mean (SD) | 264979.2 (118137.9) | 317250.0 (197420.4) | 1.00 (1.00-1.00, p=0.123) | 1.00 (1.00-1.00, p=0.818) |
| Comorbidity | No | 16 (64.0) | 9 (36.0) | - | - |
|  | Yes | 33 (43.4) | 43 (56.6) | 2.32 (0.93-6.10, p=0.078) | - |
| Pulmonary disease | No | 43 (53.1) | 38 (46.9) | - | - |
|  | Yes | 6 (30.0) | 14 (70.0) | 2.64 (0.96-8.09, p=0.070) | 1.41 (0.32-6.34, p=0.644) |
| Hypertension | No | 29 (46.8) | 33 (53.2) | - | - |
|  | Yes | 20 (51.3) | 19 (48.7) | 0.83 (0.37-1.86, p=0.66) | - |
| Cardiovascular disease | No | 39 (47.6) | 43 (52.4) | - | - |
|  | Yes | 10 (52.6) | 9 (47.4) | 0.82 (0.30-2.23, p=0.69) | - |
| Renal disease | No | 42 (50.6) | 41 (49.4) | - | - |
|  | Yes | 7 (38.9) | 11 (61.1) | 1.61 (0.58-4.76, p=0.37) | - |
| Diabetes | No | 39 (47.0) | 44 (53.0) | - | - |
|  | Yes | 10 (55.6) | 8 (44.4) | 0.71 (0.25-1.97, p=0.51) | - |
| Immunocompromise | No | 47 (50.5) | 46 (49.5) | - | - |
|  | Yes | 2 (25.0) | 6 (75.0) | 3.07 (0.67-21.67, p=0.184) | 8.79 (0.77-131.36, p=0.086) |
| Cancer | No | 49 (51.0) | 47 (49.0) | - | - |
|  | Yes | 0 (0.0) | 5 (100.0) | NE | - |
| Obesity | No | 46 (46.9) | 52 (53.1) | - | - |
|  | Yes | 3 (100.0) | 0 (0.0) | NE | - |
| Neuro disorder | No | 49 (50.0) | 49 (50.0) | - | - |
|  | Yes | 0 (0.0) | 3 (100.0) | NE | - |
| Tuberculosis | No | 49 (50.5) | 48 (49.5) | - | - |
|  | Yes | 0 (0.0) | 4 (100.0) | NE | - |
| Other comorbidities | No | 45 (47.9) | 49 (52.1) | - | - |
|  | Yes | 4 (57.1) | 3 (42.9) | 0.69 (0.13-3.29, p=0.637) | - |
| Having pets | No | 47 (47.5) | 52 (52.5) | - | - |
|  | Yes | 2 (100.0) | 0 (0.0) | NE | - |
| Living near forest | No | 47 (50.5) | 46 (49.5) | - | - |
|  | Yes | 2 (25.0) | 6 (75.0) | 3.07 (0.67-21.67, p=0.184) | 1.21 (0.11-13.47, p=0.870) |
| Comorbidity burden | None | 15 (68.2) | 7 (31.8) | - | - |
|  | Single | 12 (41.4) | 17 (58.6) | 3.04 (0.97-10.16, p=0.061) | 4.04 (0.34-56.02, p=0.275) |
|  | Multiple | 22 (44.0) | 28 (56.0) | 2.73 (0.97-8.24, p=0.063) | 1.74 (0.13-25.37, p=0.675) |

NE: Not estimable.
